# Effects of a clinical decision support system and patient portal for preventing medication-related falls in older fallers: Protocol of a cluster randomized controlled trial with embedded process and economic evaluations (AD*F*ICE_IT)

**DOI:** 10.1101/2023.07.19.23292866

**Authors:** Kelly K. de Wildt, Bob van de Loo, Annemiek J. Linn, Stephanie K. Medlock, Sara S. Groos, Kim J. Ploegmakers, Lotta J. Seppala, Judith E. Bosmans, Ameen Abu-Hanna, Julia C.M. van Weert, Natasja M. van Schoor, Nathalie van der Velde, the ADFICE_IT study team

**Affiliations:** Amsterdam School of Communication Research, University of Amsterdam, Amsterdam, the Netherlands; Amsterdam Public Health research institute, Amsterdam, The Netherlands; Amsterdam UMC location Vrije Universiteit Amsterdam, Epidemiology and Data Science, De Boelelaan 1117, Amsterdam, Netherlands; Amsterdam UMC location University of Amsterdam, Internal Medicine, Section of Geriatric Medicine, Meibergdreef 9, Amsterdam, Netherlands; Amsterdam UMC location University of Amsterdam, Department of Medical Informatics, Meibergdreef 9, Amsterdam, Netherlands; Department of Health Sciences, Faculty of Science, Vrije Universiteit Amsterdam, Amsterdam Public Health Research Institute, Van der Boechorststraat 7, 1081 BT Amsterdam, The Netherlands

**Keywords:** fall prevention, fall-risk increasing drugs (FRIDs), older patients, outpatient clinic, prediction model, Clinical decision support system, patient portal, Shared Decision Making (SDM), randomized controlled trial (RCT), study protocol

## Abstract

**Background:** Falls are the leading cause of injury-related mortality and hospitalization among adults aged ≥ 65 years. An important modifiable fall-risk factor is use of fall-risk increasing drugs (FRIDs). However, deprescribing is not always attempted or performed successfully. The AD*F*ICE_IT trial evaluates the combined use of a clinical decision support system (CDSS) and a patient portal for optimizing the deprescribing of FRIDs in older fallers. The intervention aims to optimize and enhance shared decision making (SDM) and consequently prevent injurious falls and reduce healthcare-related costs.

**Methods:** A multicenter, cluster-randomized controlled trial with process evaluation will be conducted among hospitals in the Netherlands. We aim to include 856 individuals aged ≥ 65 years that visit the falls clinic due to a fall. The intervention comprises the combined use of a CDSS and a patient portal. The CDSS provides guideline-based advice with regard to deprescribing and an individual fall-risk estimation, as calculated by an embedded prediction model. The patient portal provides educational information and a summary of the patient’s consultation. Hospitals in the control arm will provide care-as-usual. Fall-calendars will be used for measuring the time to first injurious fall (primary outcome) and secondary fall outcomes during one year. Other measurements will be conducted at baseline, 3, 6, and 12 months and include quality of life, cost-effectiveness, feasibility, and shared decision-making measures. Data will be analyzed according to the intention-to-treat principle. Difference in time to injurious fall between the intervention and control group will be analyzed using multilevel Cox regression.

**Discussion:** The findings of this study will add valuable insights about how digital health informatics tools that target physicians and older adults can optimize deprescribing and support SDM. We expect the CDSS and patient portal to aid in deprescribing of FRIDs, resulting in a reduction in falls and related injuries.

**Trial registration:** ClinicalTrials.gov NCT05449470 (7-7-2022)

**Participant recruitment:** 7 July 2022-ongoing *

* Results of this study have not yet been published or submitted to any journal.

**Protocol version:** 1

**Trial sponsor:** Amsterdam UMC, Meibergdreef 9, 1105 AZ Amsterdam

## Background

Falling among adults aged 65 years and older represents a serious public health problem. Approximately 30% of adults aged 65 or older falls each year. Moreover, falls are the leading cause of injury-related mortality and hospitalization, with one out of five falls resulting in severe injury (1). In the Western European region, 8.4 million adults aged 70 and older sought medical attention due to a fall-related injury, and 54 504 older adults died due to falls in 2017 alone (2). The incidence rate of fall-related injuries increases substantially with age (2).

Besides physical injuries such as head wounds and fractures (3,4), falls can also lead to the development of fear of falling (5,6), reduced perceived quality of life (7), reduced physical activity (8), physical decline (9), social isolation (10), increased healthcare utilization, and institutionalization (9,11,12). Furthermore, falls pose a substantial economic burden as fall-related costs are estimated to amount to 0.85 to 1.5 percent of the total healthcare expenditures in Western countries (13).

Falls have a complex etiology and are associated with several risk factors, such as history of falls (14), impaired mobility (14), frailty (15), chronic health conditions (16), fear of falling (17), depression (18), cognitive impairment (19), increasing age (18), and female gender (18). In addition, a large body of research has linked the use of certain medications to falls (20–22). Medications recognized as fall-risk increasing drugs (FRIDs) include antipsychotics, antidepressants, diuretics, and opioids (23). Studies have reported that 65 to 93 percent of older adults admitted with fall-related injuries use at least one FRID (24). Antidepressants were the most commonly used FRID at the time of the fall-related injury, with a prevalence between 15 and 40 percent (24).

Despite the growing evidence on medication as an important modifiable risk factor, deprescribing in older adults is often not attempted or performed unsuccessfully. Physicians generally find deprescribing challenging since it requires complex decision-making in the context of polypharmacy and multi-morbidity (25). To be precise, physicians find it difficult to identify which patients are at risk of a medication-related fall and it is not always clear which medications should be considered for withdrawal and whether safer alternatives are available. Moreover, patients’ beliefs regarding their medication use may further hinder effective FRIDs deprescribing. Research indicates patients are generally not concerned about possible adverse effects from their regular medication and not aware of medication management as an effective fall-prevention strategy (26,27). More effective communication may help raise awareness and consequently prompt patients to adopt to and comply with deprescribing as a treatment option. Moreover, communication is a two-way process and research suggests that interventions targeting both physicians and patients may be more effective than interventions that only target either one (28). Given these multifaceted complications, a multicomponent intervention is expected to improve FRIDs deprescribing in older adults and thereby help prevent medication-related falls.

There is growing attention for the role of SDM in deprescribing (29–31). SDM can be defined as an approach where clinicians and patients share the best available evidence when making decisions. In doing so, patients are supported to consider options and to achieve informed preferences (32). In complex patient cases with multiple treatment options, as is often the case in deprescribing in older adults, SDM has been found to lead to more informed decision-making, better participation in decision-making, more self-efficacy, increased knowledge, and reduced decisional conflict of patients in disadvantaged groups, such as older patients (33–35). Therefore, it is expected that enhanced SDM would support the FRIDs deprescribing process as well as improve patient compliance and adherence to the new treatment plan. This, in turn, may lead to a decrease in medication-related falls among older adults.

Clinical decision support systems (CDSS) may help physicians in the deprescribing process of FRIDs and may stimulate SDM. A CDSS is a computerized system that aims to support clinical-decision making by generating assessments or recommendations based on the characteristics of an individual patient. CDSSs generate patient-specific output based on an existing knowledge base or based on predictive modelling methods. CDSSs are increasingly used for improving adherence to clinical guidelines as well as for preventing prescription errors and checking for drug interactions (36). Use of CDSSs in the prevention of falls has been studied in in- and outpatient settings (37–40). However, these studies were all limited in scope as they focused on a select number of FRIDs, did not use utilize predictive modelling methods for generating patient-specific output, or did not address risk communication or shared decision-making (SDM) (37–40).

A tool that could stimulate patients to participate in SDM is a patient portal, which allows patients to access their clinical data through a secure website (41). A recent systematic literature review on the impact of patient portals on health outcomes found that patient portals can enhance preventive behaviors and adherence to therapy (42). Furthermore, a qualitative study revealed that patients thought that a portal would facilitate them in seeking medical advice in between visits (e.g., on medication side effects) and that this would stimulate patient-driven communication (43).

Given this backdrop, the AD*F*ICE_IT project (Alerting on adverse Drug reactions: Falls prevention Improvement through developing a Computerized clinical support system: Effectiveness of Individualized medicaTion withdrawal) was initiated to develop and evaluate a multicomponent intervention for optimizing FRIDs deprescribing and consequently improve patient outcomes. The intervention comprises the combined use of a CDSS and a patient portal. The CDSS includes a personalized fall risk prediction, which is used to estimate and visualizes a patient’s fall risk. Furthermore, the CDSS gives insight in which of the patient’s medications can contribute to this fall-risk, provides suggestions with safer medication alternatives, provides guideline-based medication advice, and provides an overview of the possible treatment actions. The patient portal provides general fall-related educational information (e.g. information about falls prevention, FRIDs, and FRIDs deprescribing) and information to help patients prepare for their visit to the falls clinic. After the falls clinic visit, the patient portal will show a summary of the patient’s treatment plan as discussed during the consultation. These features of the CDSS and patient portal may help to optimize and enhance (shared) decision making during the consultation. Consequently, it is expected that this will lead to less injurious falls among older adults and reduce healthcare-related costs.

The primary aim of the AD*F*ICE_IT cluster randomized controlled trial is to assess the effectiveness of the multicomponent intervention, comprised of a CDSS and patient portal, compared with usual care. Effectiveness will be assessed in terms of time to first injurious fall (primary outcome). In addition, as secondary aims we will study the cost-effectiveness and feasibility of the intervention.

## Methods

The SPIRIT criteria were used as guideline for the reporting of this protocol paper (44) (Supplementary File 1). The *CONSORT 2010 Statement: extension to cluster randomised controlled trials* will be used to further guide the reporting of the results of the trial (45).

The design and the development of the AD*F*ICE_IT intervention was guided by the Medical Research Council (MRC) Framework for Complex Interventions (46). In the preparation phase of the MRC framework, we developed a prediction model for estimating a patient’s risk of falling (47). The prediction model is currently being externally validated. In the development phase, we identified evidence and theory regarding CDSS and patient portal end users’ preferences and needs, and extended these with empirical research (i.e. survey (48)), interviews) to inform our decisions regarding the design of the intervention. Furthermore, we incorporated guideline- and expert consensus-based medication advices (e.g. deprescribing advice or use of safer alternative medication) in the CDSS (23). In the feasibility/piloting phase of the MRC framework, we tested the usability of the user interface of our intervention through usability studies. The present paper describes the protocol for the final phase of the project in which we will evaluate the effectiveness of the intervention.

### Study design and settings

To evaluate the effectiveness of our multicomponent (CDSS and patient portal) intervention in preventing injurious falls among older adults, a multicenter cluster-randomized controlled trial will be conducted among new falls clinic patients of ten Dutch hospitals. These patients have been referred for a multifactorial falls assessment to the geriatrics departments by their general practitioner, the emergency department, or other specialists because of a history of falling or an increased risk of falling.

### Ethical considerations

The ADFICE_IT study protocol was reviewed and approved by the Medical Ethics review board of the Amsterdam University Medical Centres (METC AMC 2021_061). All study participants will asked to sign an informed consent prior to data collection. The trial is registered with ClinicalTrials.gov (DATE; 7-7-2022, identifier: NCT05449470).

### Eligibility criteria

The study population consists of older adults visiting a falls clinic. Falls clinics typically perform detailed multidisciplinary fall risk assessments and make recommendations or implement a range of targeted falls and falls injury-prevention strategies based on the assessment findings (49). Falls clinics at Dutch hospitals that use Epic software (Epic Systems Corporation; Verona, Wisconsin, United States) as their electronic patient record system were eligible to be included as a study center. Patients meeting the following criteria are eligible for inclusion:

– Aged 65 years and older;
– History of at least one fall in the past year;
– A Mini-Mental State Examination (MMSE) score of 21 points or higher or equivalently a Montreal Cognitive Assessment (MOCA) Dutch score of 16 points or higher (50);
– Use of at least one FRID (as defined by the Dutch Federation of Medical Specialists (51));
– Sufficient command of the Dutch language in speech and writing; and
– Willingness to sign informed consent.

Potential subjects will be excluded if they:

– Already participate in another (intervention) study;
– Have a life expectancy of less than one year; or
– Suffer from severe mobility impairment (i.e. bedridden, e.g. inability to walk short distances with assistance of a walking aid).

Participant recruitment has started in July 2022 and is ongoing.

### Randomization and blinding

Since the intervention needs to be integrated into the physician’s workflow, randomization will be performed at hospital level prior to the start of inclusion. We evaluated use of the CDSS in usability studies among physicians of one of the locations of the Amsterdam UMC, i.e. location AMC. To avoid possible contamination of the intervention, the Amsterdam UMC: location AMC will be exempted from randomization and included in the intervention group by default. To assure the control and intervention hospitals remain similar with respect to their patient population, the other location of Amsterdam UMC, i.e. location VUmc, will be included in the control group by default. Randomization of the remaining hospitals will be done based on a 1:1 allocation ratio and stratified based on whether the hospital is academic or non-academic. The randomization procedure will be done by an independent statistician using computer-generated random numbers. Blinding of the intervention allocation is not possible since both physicians and patients will have to interact with the CDSS and patient portal. Researchers will be blinded to group allocation during the statistical analyses.

### Intervention

The multicomponent intervention comprises the combined use of a CDSS and a patient portal. Furthermore, patients in the intervention arm will receive a Question Prompt List (QPL) prior to their consultation. Physicians in the intervention arm will be trained to work with the CDSS. The control hospitals will only receive a general overview of the study, including the procedures. Patients in the control arm will receive care-as-usual.

#### CDSS

Relevant FRIDs were identified based on the Dutch fall guideline (51) and STOPPFall (Screening Tool of Older Persons Prescriptions in older adults with high fall risk) tool (23). These two sources form the foundation for the CDSS’ clinical knowledge base. For each class of identified FRIDs, relevant recommendations about deprescribing from more than 30 different Dutch clinical guidelines have been extracted and formalized using the Logical Elements Rule Method (52). Thus, the CDSS provides point-of-care guideline and expert consensus based medication withdrawal advice (23) as well as a personalized fall-risk estimation based on a prediction model (47).

The CDSS will be integrated in the electronic patient record system and workflow of physicians. On the CDSS start page, the physician can check the data that was pulled from the electronic patient record system, and see the patient’s estimated risk of falling. On the next screen, the physician can see the advice of the CDSS for each of the patient’s prescribed current medications. Based on the given advice, the physician can decide to propose a change in treatment for a specific medication. The physician can discuss those proposed treatment changes with the patient using the consultation screen. The final screen will allow the physician to copy-paste all treatment decisions to the patient’s electronic health record, print a patient-friendly summary of the individual treatment plan, and send it to the patient portal.

#### Patient portal

Patients in the intervention arm will receive access to the patient portal prior to their visit to the falls clinic. At that time, the patient portal provides general fall-related educational information (e.g. information about falls prevention, FRIDs, and FRIDs deprescribing) and information to help patients prepare for the falls clinic visit. After their consultation with the physician during their fall clinic visit, the patients will also receive access to the additional patient portal pages with the personalized fall-risk estimate and the treatment plan as discussed with the physician.

#### Question Prompt List (QPL)

Patients in the intervention arm will receive a printed QPL prior to their visit to the falls clinic. A QPL is a structured list of questions designed to encourage information gathering, which patients can use as example questions to ask during the consultation (53). A QPL stimulates agenda setting and helps patients to remember important questions. In other contexts (e.g. oncology), QPLs have been found to improve communication and stimulate participation in older patients (53). Our QPL will consist of preparatory questions and concerns that need to be completed by the patient preceding the consultation (e.g. ‘Which of the medications that I am currently taking are truly crucial for my health?’). Patients will be asked to bring it with them to consultation.

#### Training

Physicians will be trained in small groups (i.e., the geriatric staff of a specific intervention hospital) on how to use the system during a one-hour training session. The training addresses four components: 1) general overview of the study and its aim, 2) (issues in) FRIDs deprescribing, 3) employing SDM and the QPL during a consultation, and 4) practical instructions on how to use the CDSS. AD*F*ICE_IT project team members (i.e., an experienced geriatrician and two communication scholars) will provide the training. Afterwards, an online version of the training will be available to the physicians.

### Comparator

Patients treated at the control hospitals will receive care-as-usual, e.g. a multifactorial fall assessment at a falls clinic. Usually such an assessment takes up around 3-4 hours, distributed over 1 or 2 days, and is concluded by a consultation between the patient and the physician.

### Procedures

Patients who schedule an appointment at any of the participating falls clinics (i.e., intervention and control hospitals) will receive a letter containing information on the objectives and procedures of the study and an invitation to participate. For patients in the intervention arm, the invitation letter will also include a printed QPL and a link to the patient portal.

At the falls clinic, eligibility will be determined according to the in- and exclusion criteria by the hospitals’ staff members. The researcher will then provide oral and written information about the study to eligible patients. Patients who are interested in participating in the study will be asked to sign an informed consent form. Next, the falls clinic assessments will be carried out as usual. In the intervention group, the physician will use the CDSS prior to the consultation to understand a patient’s fall risk and medical background as well as during the consultation with the patient. Consultations in the control group are carried out according to care as usual. After the consultation, the research assistants will ask the included patients and their caregivers (if applicable) to fill out a set of questionnaires (see “Data Collection”). After the visit to the falls clinic, patients in the intervention group will be able to review information about their consultation with the physician (i.e., their treatment plan) and their estimated fall risk in the patient portal.

### Data collection

We will collect a wide range of quantitative and qualitative data to assess the effectiveness and cost-effectiveness and to evaluate the implementation of the intervention (see Figure 1 for complete overview of measurements). Questionnaires will be administered at baseline and 3, 6, and 12 months after baseline (Figure 1).

**Figure 1.**
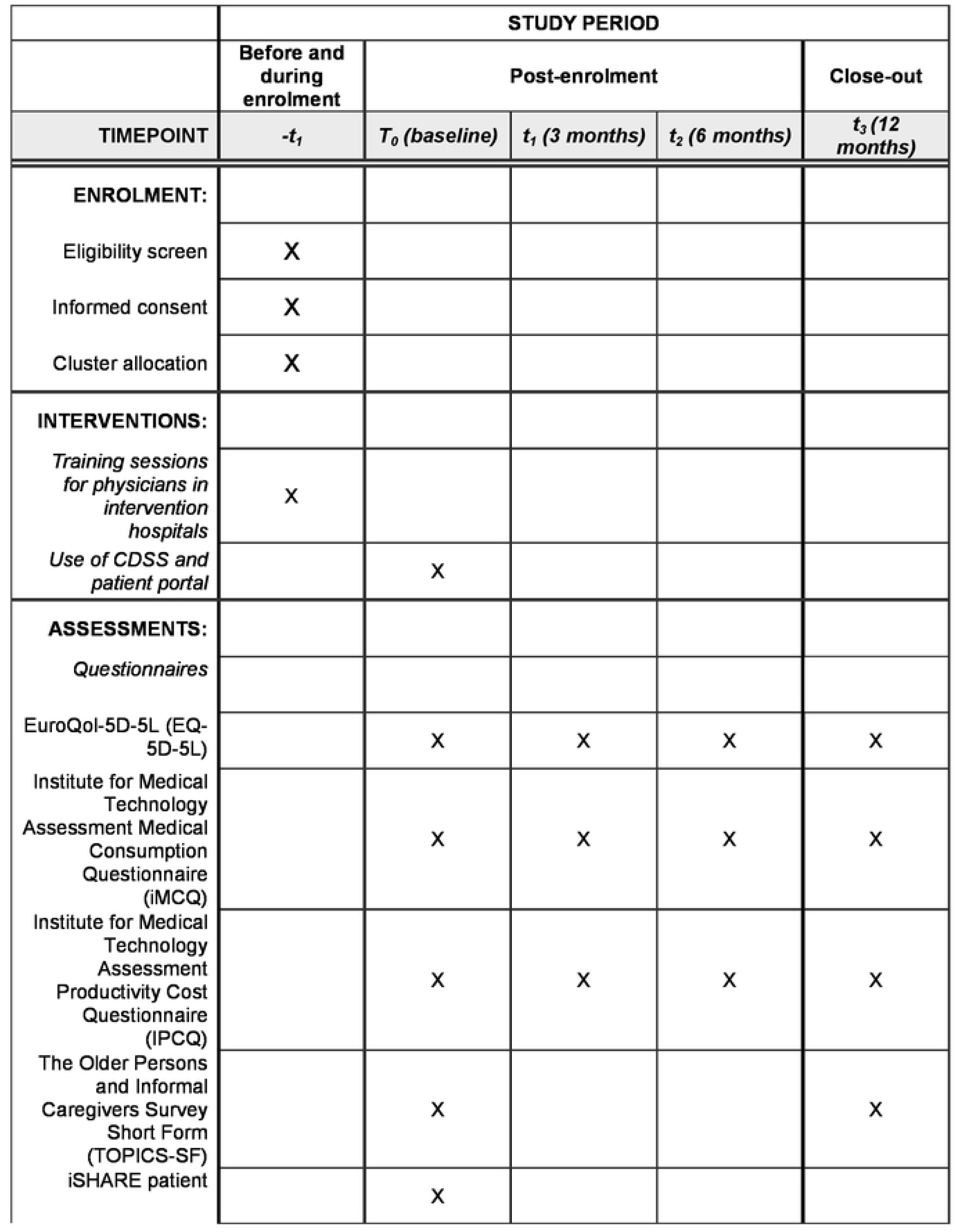
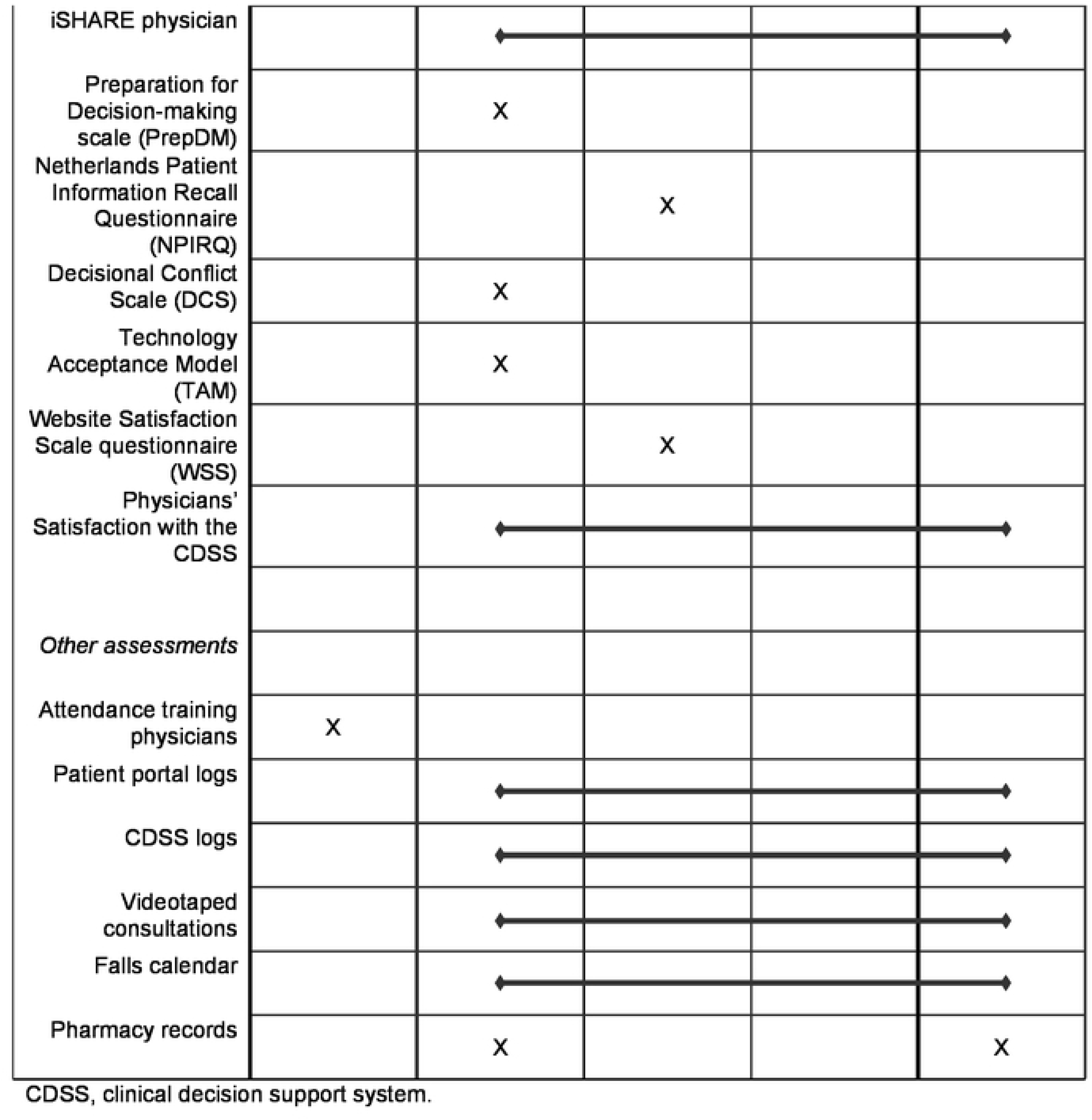
Schedule of study procedures and assessments.

### Estimating the effectiveness of the intervention on trial outcomes

The primary outcome is time to first injurious fall. An injurious fall is defined as a fall resulting in wounds, bruises, sprains, cuts, medically recorded fractures, head or internal injury, requiring medical/health professional examination, accident and emergency treatment, or inpatient treatment (54). This definition is consistent with moderate and serious injuries, as proposed by Schwenk (2012).

Secondary outcomes include number of injurious falls, total number of falls, time to first fall resulting in any injuries (i.e., fall that results in minor, moderate, or severe injuries), total number of falls resulting in any injuries, time to first fall and (health-related) quality of life. Falls are defined as an unexpected event in which the participants come to rest on the ground, floor, or lower level (55). At baseline, patients in both groups receive a falls calendar to keep track of falls, fall-related injuries, and fall-related healthcare use on a weekly basis for 12 months. The falls calendars will be returned every month by mail. Incomplete, missing or unclear data will be further inquired by telephone. Health-related) quality of life is assessed using the EuroQol-5D-5L (EQ-5D-5L) index value, EQ-5D visual analogue scale and the Older Persons and Informal Caregivers Survey Short Form (TOPICS-SF) summary score (56,57). EQ-5D-5L is a standardized instrument for measuring health-related quality of life (58). The health states based on the five EQ-5D-5L domains will be converted to utility scores using the Dutch EQ-5D-5L tariff (56). The TOPICS-SF is a 22-item questionnaire for measuring health-related quality of life, which was developed to evaluate patient-reported outcomes in the context of multidimensional geriatric care (57).

### Estimating the cost-effectiveness of the intervention

Societal costs related to the intervention and care as usual will be assessed using the institute for Medical Technology Assessment Medical Consumption Questionnaire (iMCQ) (59) and the institute for Medical Technology Assessment Productivity Cost Questionnaire (iPCQ; Figure 1) (60). The iMCQ is a non-disease specific questionnaire for measuring health care use (59). The iPCQ is a questionnaire for measuring productivity losses of paid work due to absenteeism, presenteeism and productivity losses related to unpaid work (60). Costs will be calculated by multiplying the volumes of healthcare use with the corresponding unit prices. Lost productivity costs will be calculated using the friction cost approach.

### Process evaluation

The process evaluation will consist of two parts: a) assessing the feasibility of the intervention and b) evaluating how the intervention facilitates SDM.

#### Process evaluation: Feasibility

To assess the feasibility of the intervention, we will collect the following: 1) data logged by the CDSS and patient portal, 2) participation data of the CDSS training, 3) physician satisfaction regarding the CDSS, 4) the Technology Acceptance Model (TAM; (61)) questionnaire; 5) the Website Satisfaction Scale questionnaire (WSS (62)), 6) videotaped consultations, 7) pharmacy records, and 8) falls calendar entries.

Usage data of the CDSS and patient portal will be measured throughout the study period. The extent to which the intervention was implemented as intended (fidelity/dose delivered) and the extent to which the participants actively engage with the intervention (dose received/exposure) will be assessed through data logged by the CDSS and patient portal, and the videotaped consultations. Dose received/exposure will also be assessed through participation data of the CDSS training. Reach/participation rate will be assessed through data logged by the CDSS to analyze the extent to which physicians propose changes in FRID prescriptions. Patients will be asked to self-report changes in medication use on the fall calendar on a weekly basis. These falls calendar entries and pharmacy records will be used to analyze the extent to which patients adhere to the physicians’ advice and changes in the treatment plan. Satisfaction with the intervention (dose received/exposure) will be measured through physician evaluations of the CDSS and the WSS questionnaire (patients). The TAM questionnaire assesses the perceived usefulness, perceived ease of use, and intended usage of the CDSS, and the WSS measures the comprehensibility, satisfaction and Emotional Support of the patient portal (62). Finally, barriers and facilitators (context) will be assessed through the videotaped consultations, and physician evaluations of the CDSS by means of a survey.

#### Process evaluation: Shared Decision Making

SDM will be measured through self-reported questionnaires in the full sample (i.e. perceived SDM; the iSHAREpatient and iSHAREphysician (63). In addition, we aim to measure observed SDM in a subsample (n=50) through videos of consultations (i.e. the Observer OPTION^MCC^ Multiple Chronic Conditions coding scheme (64)). SDM will be assessed in relation to two affective-cognitive outcomes: preparation for decision-making through the Preparation for Decision-making scale (PrepDM; (65)) and decisional conflict through the Decisional Conflict Scale (DCS; low literacy version; (66)). The ‘Question Format DCS – 10 item 3 response categories’ version of the DCS is recommended to be used for low literacy groups (67). In addition, recall of information will be assessed in the subsample through the Netherlands Patient Information Recall Questionnaire (NPIRQ; (68)). The iSHARE questionnaire will be used to measure perceived SDM from both the patient and physician perspective. The NPIRQ consists of multiple-choice questions, completion items, and open-ended questions related to information about treatment and recommendations on how to handle side effects (68). Patient responses on the questionnaire will be checked against the actual communication in video recordings of the consultations. In addition, the PrepDM will be used to assess how patients evaluate the usefulness of the patient portal and QPL for preparing themselves for communicating with their physician during the consultation. Finally, we will code observed shared (triadic) decision-making between the physician, the patient, and if relevant, the informal caregiver, in the videotaped consultations by using the Observer OPTION^MCC^ Multiple Chronic Conditions coding scheme (64).

SDM will be assessed using questionnaires in the full sample and video observations in a subsample. The subsample of 50 consultations in both intervention and control group will be video recorded to assess the level of SDM. After working with the CDSS for a couple of months, intervention-group physicians will be asked to fill out an online questionnaire about their satisfaction with the CDSS, to indicate whether they thought the advice provided by the CDSS was (sufficiently) accurate, if they perceived any barriers in using the CDSS system, and if they thought the patient perceived barriers in using the patient portal. Pharmacy records will be used to make an inventory of the prescribed medicines for individual patients at baseline and 12 months after baseline to assess adherence.

### Data management

Data will be handled confidentially and only a limited number of members of the study team will have access to the complete datasets. The collected and pseudonymized questionnaire data for each local center will be transferred to the Amsterdam UMC, where it will be entered, stored and processed in Castor. In addition, the digital CDSS and patient portal data will be stored locally at each hospital. Every 3-6 months, study data will be extracted to .csv text files and stored in a secured folder.

Furthermore, administrative data will be stored in a secured SQL database. Finally, data from both control and intervention patients will be extracted from Epic every 3-6 months to .csv text files (e.g. medication data, problem lists, relevant lab values, and the prediction model variables). Data from individual patients will be pseudonymized, and the different datasets can only be linked through a participant identification number, which is stored in a separate data system. These data management systems all comply in accordance with the European Union General Data Protection Regulation.

### Statistical analysis

Data of the RCT will be analyzed according to the intention-to-treat principle. P-values of < 0.05 will be considered statistically significant.

#### Estimating the effectiveness of the intervention on trial outcomes

For every participant, we will assess fall incidents during a fixed follow-up period of 12 months, which will start after a set 1 month, during which the dose of FRIDs will be stopped or decreased. Difference in time to injurious fall between the intervention and control group in the follow-up period will be analyzed by means of a multilevel Cox regression model based on hospital level (69). Model fit will be assessed using standard approaches (e.g., the proportional hazards assumption with Schoenfeld residuals). We will adjust all models for age, sex and type of hospital, i.e. academic versus non-academic. In a sensitivity analysis, we will additionally adjust for significant baseline differences. Difference in total number of (injurious) falls in the follow-up period between the control and intervention groups will be analyzed by means of multilevel Poisson regression models based on hospital level. In the case of overdispersion, we will apply either quasi-Poisson regression or negative binomial regression depending on the observed distribution of the data. Difference between the intervention and control group with respect to time to any fall and time to fall that results in any injuries will be analyzed by means of survival analyses, similarly to the primary outcome.

Differences in EQ-5D-5L index score, EQ-5D visual analogue scale, and TOPICS-SF summary score between the intervention and control group after 12 months will be analyzed by means of linear mixed models. These models will be adjusted for the baseline value of the outcome (70).

#### Estimating the cost-effectiveness of the intervention

Differences in costs and effects between intervention and usual care will be estimated using seemingly unrelated regression to retain the correlation between costs and effects. Incremental cost-effectiveness ratios will be calculated by dividing the difference in costs between CDSS and usual care due to differences in incidence in injurious falls as well as gained quality-adjusted life years (QALYs).

Bootstrapping techniques will be used to estimate the uncertainty surrounding the incremental cost-effectiveness ratios. Uncertainty will be shown in cost-effectiveness planes and cost-effectiveness acceptability curves.

#### Process evaluation: Feasibility

In the first part of the process evaluation we will evaluate the feasibility of the intervention, and describe 1) user data, 2) participation in and evaluation of the training, 3) physician and patient satisfaction and acceptance of the CDSS and patient portal. These descriptive statistics will be presented as percentages or means with standard deviations.

#### Process evaluation: Shared Decision Making

Differences in mean change between arms will be analyzed with the use of multi-level modelling and will be expressed as mean differences with 95% CIs. We will analyze differences in perceived SDM and observed SDM for both patients and physicians in the intervention and control groups. Finally, we will assess the differences between the intervention and control groups on recall (NPIRQ), adherence (pharmacy records), and quality of life (EQ-5D-5L).

### Per protocol analysis

Logged data by the CDSS and patient portal will be used to select participants for a per protocol analysis for the primary outcome. In this analysis, we will only include patients from the experimental group that meet the following two criteria 1) physicians used the CDSS in the consult with the patients and 2) physician used the ‘print’ button in the CDSS or the patient visited the patient portal at least once after the consultation.

### Sample size

Sample sizes of *n* = 385 in the intervention and *n* = 385 in the control group (10 clusters with 77 patients in each cluster) are needed to detect a difference in proportion of injurious falls of 0.10 with 80% power. We inferred the proportion of patients who will experience an injurious fall to be 0.22 in the control and 0.12 in the intervention group. In these calculations, we assumed the two-sided significance level of 5% and an intraclass correlation coefficient of 0.01 to account for clustering (71). Presupposing a drop-out rate of 10%, 856 patients will need to be included.

### Monitoring

A data monitoring committee will not be established since the overall risk associated with the trial is considered negligible.

### Harms

All adverse events and serious adverse events reported by the subject or observed by the researchers or his staff will be recorded in an electronic database. Serious adverse events will also be reported to the medical ethics committee of the Amsterdam UMC.

## Discussion

The multicenter RCT described in this paper will assess: a) the effectiveness of the multicomponent intervention (i.e., use of CDSS and patient portal) compared with usual care. Effectiveness will be assessed in terms of time to first injurious fall (primary outcome). As secondary aims, cost-effectiveness and the feasibility of the intervention will be assessed.

The deprescribing of FRIDs requires complex decision making. We expect that the implementation of our CDSS and patient portal, supported by a prediction model and guideline-based advice, will aid in optimizing deprescribing decisions for both the physician and patient, consequently reducing fall risk. In line with the expectation that the intervention will aid in the prevention of injurious falls, it is hypothesized that the intervention will be more cost-effective compared to care-as-usual regarding fall-related health care costs. The direct healthcare and follow-up care resulting from injurious falls among older adults potentially involve 0.85 to 1.5 percent of the total healthcare expenditures in Western countries (13).

The process evaluation will evaluate a) the implementation of the intervention and b) how this intervention leads to enhanced SDM and patient outcomes. A systematic review has suggested that SDM can lead to better affective-cognitive outcomes, e.g. improved satisfaction and less decisional conflict (72). Thus, we hypothesize that physicians and geriatric patients as well as their caregivers will evaluate the intervention workflow more positively compared to the care-as-usual workflow and will engage in more SDM regarding the patient’s treatment plan. Recent studies have illustrated that compliance to FRID-deprescribing is often poor. In a study by Boyé et al. (73), researchers found that compliance to their intervention of FRIDs-withdrawal was limited among patients. The researchers found that 35 percent of all deprescribing attempts were unsuccessful, either due to non-compliance, recurrence of the initial indication for prescribing, or additional medication being described for newly diagnosed conditions. Moreover, the STRIDE trial, a multicenter randomized controlled trial by Basin et al. (74) evaluated a multifactorial intervention that included the use of motivational interviewing to encourage patients to choose recommendations they were willing to address. Among the patients for which medication use was identified as a risk factor, only 29 percent of patients agreed to address this risk. We expect that a higher degree of SDM will lead to more recall and knowledge among patients, leading to more treatment and medication adherence among patients. This in turn, could also lead to less medication-related injurious falls among patients.

An important strength of our study is that we developed the intervention following the MRC guidelines. The aim of the framework is to ensure that feasible interventions are empirically and theoretically founded and that considerations are given both to the effectiveness of the intervention and how it works. The intervention’s end-users are included in each phase of the project. This way, we will be able to optimally personalize the intervention’s design to the heterogeneous needs of the end-users.

Another asset of our study is that it includes both an effect evaluation and a process evaluation. This will help us to not only assess whether the intervention was effective, but the process evaluation will also make it possible to assess whether the intervention was implemented correctly, and which implementation factors were facilitating or impeding. Gaining more insight into the context will deepen our understanding of why the intervention was (not) successful.

The findings of this study will add valuable insights about how digital health informatics tools, based on prediction models, can support SDM between physicians and older adults. This new knowledge will be especially insightful in the case of FRIDs withdrawal among older adults.

Furthermore, this study will also contribute to the literature on risk communication, since it investigates how physicians will use a visualized fall-risk estimate in their consultations with the patient.

## Outlook

If the AD*F*ICE_IT intervention will prove to be effective, it could be implemented in routine healthcare practices. The hospitals in the intervention group can continue using the CDSS and patient portal as they have done during the RCT, as the intervention will already be implemented in their electronic patient record systems. The hospitals in the control group could also implement the CDSS and patient portal software at the end of the study. Furthermore, the software will be available as open source to facilitate national and international implementation. We expect that once implemented at the falls clinic of the geriatric departments, the AD*F*ICE_IT intervention will contribute to individualized and cost-effective prevention of (medication-related) injurious falls among older adults.

## Abbreviations

AD*F*ICE_IT: Alerting on adverse Drug reactions: Falls prevention Improvement through developing a Computerized clinical support system: Effectiveness of Individualized medicaTion withdrawal

CDSS: Clinical decision support system

CI: confidence interval

DCS: Decisional Conflict Scale

EQ-5D-5L: EuroQol-5D-5L

FRID: Fall-Risk Increasing Drug

IMCQ: institute for Medical Technology Assessment Productivity Cost Questionnaire Medical Consumption Questionnaire

IPCQ: institute for Medical Technology Assessment Productivity Cost Questionnaire Productivity Cost Questionnaire

MMSE: Mini-Mental State Examination

MOCA: Montreal Cognitive Assessment

MRC: Medical Research Council

NPIRQ: Netherlands Patient Information Recall Questionnaire

PrepDM: Preparation for Decision-making scale

QALY: quality-adjusted life years

QPL: Question Prompt List

RCT: Randomized Controlled Trial

SDM: Shared Decision Making

TAM: Technology Acceptance Model

TOPICS-SF: The Older Persons and Informal Caregivers Survey – Short Form

UI: User Interface

WSS: Website Satisfaction Scale

## Dissemination policy

The results of the AD*F*ICE_IT study will be published in international, peer-reviewed, scientific journals. In addition, results will be presented at (inter)national scientific conferences, seminars, public events, and on the project website.

## Consent for publication

Not applicable.

## Availability of data and materials

All project data will be stored in accordance with General Data Protection Regulation. Data from the trial will be made available for other researchers after the study is completed for replication purposes and for original research questions. To obtain data, researchers will need to submit an analysis proposal, which will be evaluated by the AD*F*ICE_IT Steering Group. More information on the AD*F*ICE_IT study can be found at http://www.onderzoeknaarvallen.nl (website information is also available in English).

## Authors’ contributions

NvdV, NMvS, AA-H, SM, JCMvW contributed in the conception of the study. All authors have made substantial contributions to the design of the study. BvdL and KKdW wrote the manuscript. All the authors read the draft, made contributions and approved the final manuscript.

## Data Availability

Data from the trial will be made available for other researchers after the study is completed for replication purposes and for original research questions. To obtain data, researchers will need to submit an analysis proposal, which will be evaluated by the ADFICE_IT Steering Group. More information regarding the data will be made available on our study website: http://www.onderzoeknaarvallen.nl (website information is also available in English).

## Acknowledgements

Not applicable

## Authors’ information (optional)

Not applicable

## Items from the World Health Organization Trial Registration Data Set

1. Primary Registry and Trial Identifying Number Name of Primary Registry, and the unique ID number assigned by the Primary Registry to this trial. Name of Primary Registry: ClinicalTrials.gov ID number assigned by the Primary Registry to this trial: NCT05449470
Date of Registration in Primary Registry Date when trial was officially registered in the Primary Registry. 7-7-2022
Secondary Identifying Numbers

Other identifiers besides the Trial Identifying Number allocated by the Primary Registry, if any. These include:

- The Universal Trial Number (UTN)
- Identifiers assigned by the sponsor (record Sponsor name and Sponsor-issued trial number (e.g. protocol number))
- Other trial registration numbers issued by other Registries (both Primary and Partner Registries in the WHRegistry Network, and other registries)
- Identifiers issued by funding bodies, collaborative research groups, regulatory authorities, ethics committees / institutional review boards, etc.

All secondary identifiers will have 2 elements: an identifier for the issuing authority (e.g. NCT, ISRCTN, ACTRN) plus a number.

There is nlimit tthe number of secondary identifiers that can be provided.

Identifier as given by the Dutch Central Committee on Research Involving Human Subjects (https://english.ccmo.nl/): NL76386.018.21

Identifier as given by the Medical Ethics review board of the Amsterdam University Medical Centres: METC AMC 2021_061

1. Source(s) of Monetary or Material Support

Major source(s) of monetary or material support for the trial (e.g. funding agency, foundation, company, institution).

The ADFICE_IT study is supported by funding from the Netherlands Organization for Health Research and Development (ZonMw, Grant 848017004), The Hague and the Amsterdams Universiteitsfonds: Gepersonaliseerde Medicatieaanpassing bij Oudere Vallers.

5. Primary Sponsor

The individual, organization, group or other legal entity which takes responsibility for initiating, managing and/or financing a study. The Primary Sponsor is responsible for ensuring that the trial is properly registered. The Primary Sponsor may or may not be the main funder.

The sponsor of this study is the Amsterdam UMC.

6. Secondary Sponsor(s)

Additional individuals, organizations or other legal persons, if any, that have agreed with the primary sponsor ttake on responsibilities of sponsorship.

A secondary sponsor may have agreed to:

take on all the responsibilities of sponsorship jointly with the primary sponsor; or
form a group with the Primary Sponsor in which the responsibilities of sponsorship are allocated among the members of the group; or
act as the Primary Sponsor’s legal representative in relation tsome or all of the trial sites.

There are nsecondary sponsors.

1. Contact for Public Queries

Email address, telephone number and postal address of the contact whwill respond tgeneral queries, including information about current recruitment status.

“Note: The information provided in here is functional and not personal, it is recommended tprovide institutional and not personal information. By providing this information the registrant consents that the information provided can or may be published on a public website. Once provided the information cannot be redacted or anonymized as a result of new privacy legislation such as the European General Data Protection Regulation (GDPR)”.

Contact: N. van der Velde Telephone number: +3120 566 9111

Address: Amsterdam UMC location University of Amsterdam, Internal Medicine, Section of Geriatric Medicine, Meibergdreef 9, 1105 AZ Amsterdam, Netherlands

1. Contact for Scientific Queries

There must be clearly assigned responsibility for scientific leadership ta named Principal Investigator. The PI may delegate responsibility for dealing with scientific enquiries ta scientific contact for the trial. This scientific contact will be listed in addition tthe PI.

“Note: The information provided in here is functional and not personal, it is recommended tprovide institutional and not personal information. By providing this information the registrant consents that the information provided can or may be published on a public website. Once provided the information cannot be redacted or anonymized as a result of new privacy legislation such as the European General Data Protection Regulation (GDPR)”.

The contact for scientific queries must include:

- Name and title, email address, telephone number, postal address and affiliation of the Principal Investigator, and;
- Email address, telephone number, postal address and affiliation of the contact for scientific queries about the trial (if applicable). The details for the scientific contact may be generic (that is, there does not need tbe a named individual): e.g. a generic email address for research team members qualified tanswer scientific queries.

Contact: Prof. Dr. Nathalie van der Velde Telephone number: +3120 566 9111

Address: Amsterdam UMC location University of Amsterdam, Internal Medicine, Section of Geriatric Medicine, Meibergdreef 9, 1105 AZ Amsterdam, Netherlands

9. Public Title

Title intended for the lay public in easily understood language.

A Clinical Decision Support System and Patient Portal for Preventing Medication-related Falls in Older Patients

10. Scientific Title

Scientific title of the study as it appears in the protocol submitted for funding and ethical review. Include trial acronym if available.

A Clinical Decision Support System and Patient Portal for Preventing Medication-related Falls in Older Patients (ADFICE_IT)

11. Countries of Recruitment

The countries from which participants will be, are intended tbe, or have been recruited at the time of registration.

Netherlands

1. Health Condition(s) or Problem(s) Studied

Primary health condition(s) or problem(s) studied (e.g., depression, breast cancer, medication error).

If the study is conducted in healthy human volunteers belonging tthe target population of the intervention (e.g. preventive or screening interventions), enter the particular health condition(s) or problem(s) being prevented.

Prevention of medication-related injurious falls.

13. Intervention(s)

For each arm of the trial record a brief intervention name plus an intervention description.

Intervention Name: For drugs use generic name; for other types of interventions provide a brief descriptive name.

For investigational new drugs that dnot yet have a generic name, a chemical name, company code or serial number may be used on a temporary basis. As soon as the generic name has been established, update the associated registered records accordingly.
For non-drug intervention types, provide an intervention name with sufficient detail sthat it can be distinguished from other similar interventions.

<u>Intervention Description</u>: Must be sufficiently detailed for it tbe possible tdistinguish between the arms of a study (e.g. comparison of different dosages of drug) and/or among similar interventions (e.g. comparison of multiple implantable cardiac defibrillators). For example, interventions involving drugs may include dosage form, dosage, frequency and duration.

If the intervention is one or more drugs then use the International Non-Proprietary Name for each drug if possible (not brand/trade names). For an unregistered drug, the generic name, chemical name, or company serial number is acceptable.

If the intervention consists of several separate treatments, list them all in one line separated by commas (e.g. “low-fat diet, exercise”).

For controlled trials, the identity of the control arm should be clear. The control intervention(s) is/are the interventions against which the study intervention is evaluated (e.g. placebo, ntreatment, active control). If an active control is used, be sure tenter in the name(s) of that intervention, or enter “placebo” or “ntreatment” as applicable. For each intervention, describe other intervention details as applicable (dose, duration, mode of administration, etc).

Intervention Name: AD*F*ICE_IT CDSS and Patient Portal for optimizing deprescribing of fall-risk-increasing drugs

In this study we will evaluate the effect of an intervention comprised of the combined use of a clinical decision support system and a patient portal for optimizing the deprescribing of FRIDs in older fallers. Patients in the control arm will receive care-as-usual.

14. Key Inclusion and Exclusion Criteria

Inclusion and exclusion criteria for participant selection, including age and sex. Other selection criteria may relate tclinical diagnosis and co-morbid conditions; exclusion criteria are often used tensure patient safety.

If the study is conducted in healthy human volunteers not belonging tthe target population (e.g. a preliminary safety study), enter “healthy human volunteer”.

Patients meeting the following criteria are eligible for inclusion:

- Aged 65 years and older;
- History of at least one fall in the past year;
- A Mini-Mental State Examination (MMSE) score of 21 points or higher or equivalently a Montreal Cognitive Assessment (MOCA) Dutch score of 16 points or higher (50);
- Use of at least one FRID (as defined by the Dutch Federation of Medical Specialists (51));
- Sufficient command of the Dutch language in speech and writing; and
- Willingness tsign informed consent.

Potential subjects will be excluded if they:

- Already participate in another (intervention) study;
- Have a life expectancy of less than one year; or
- Suffer from severe mobility impairment (i.e. bedridden, e.g. inability twalk short distances with assistance of a walking aid).
15. Study Type

Study type consists of:

- Type of study (interventional or observational)
- Study design including:
- Method of allocation (randomized/non-randomized)
- Masking (is masking used and, if so, whis masked)
- Assignment (single arm, parallel, crossover or factorial)
- Purpose
- Phase (if applicable)

For randomized trials: the allocation concealment mechanism and sequence generation will be documented.

This study is a a multicenter, cluster-randomized controlled trial. Masking will not be used. Assignment will be at random and at the level of the cluster.

16. Date of First Enrollment

Anticipated or actual date of enrolment of the first participant.

The first patient was enrolled on 7 July 2022.

17. Sample Size

Sample Size consists of:

Number of participants that the trial plans tenrol in total.
Number of participants that the trial has enrolled.

The trial plans on enrolling 856 participants. A total of 8 participants have currently been enrolled at the point of this submission.

18. Recruitment Status

Recruitment status of this trial:

Pending: participants are not yet being recruited or enrolled at any site
Recruiting: participants are currently being recruited and enrolled
Suspended: there is a temporary halt in recruitment and enrolment
Complete: participants are nlonger being recruited or enrolled
Other

The status of this trial is: recruiting (participants are currently being recruited and enrolled).

19. Primary Outcome(s)

Outcomes are events, variables, or experiences that are measured because it is believed that they may be influenced by the intervention.

The Primary Outcome should be the outcome used in sample size calculations, or the main outcome(s) used tdetermine the effects of the intervention(s). Most trials should have only one primary outcome.

For each primary outcome provide:

The name of the outcome (dnot use abbreviations)
The metric or method of measurement used (be as specific as possible)
The timepoint(s) of primary interest

Example:

Outcome Name: Depression

Metric/method of measurement: Beck Depression Score Timepoint: 18 weeks following end of treatment

The primary outcome is time tfirst injurious fall. Injurious falls will be recorded prospectively over a period of one year, using weekly fall calendars.

20. Key Secondary Outcomes

Secondary outcomes are outcomes which are of secondary interest or that are measured at timepoints of secondary interest. A secondary outcome may involve the same event, variable, or experience as the primary outcome, but measured at timepoints other than those of primary interest.

As for primary outcomes, for each secondary outcome provide:

The name of the outcome (dnot use abbreviations)
The metric or method of measurement used (be as specific as possible)
The timepoint(s) of interest

Number of injurious falls [Time Frame: 12 months]

This concerns the total number of injuirous falls over the course of 12 months. An injurious fall is defined as a fall resulting in wounds, bruises, sprains, cuts, medically recorded fractures, head or internal injury, requiring medical/health professional examination, accident and emergency treatment, or inpatient treatment.

Total number of falls [Time Frame: 12 months]

Total number of any fall (I.e. a fall that results in ninjuries, or minor, moderate, or severe injuries)

Time tfirst fall resulting in any injuries [Time Frame: 12 months]

I.e. a fall that results in minor, moderate, or severe injuries

Total number of falls resulting in any injuries [Time Frame: 12 months]

I.e. a fall that results in minor, moderate, or severe injuries

Time tfirst non-injurious fall [Time Frame: 12 months]

I.e. a fall that results in ninjuries

EuroQol-5D-5L (EQ-5D-5L) [Time Frame: at baseline, 3 months, 6 months, and 12 months]

The descriptive system comprises five dimensions: mobility, self-care, usual activities, pain/discomfort and anxiety/depression. Each dimension in the EQ-5D-5L has five response levels: nproblems (Level 1); slight; moderate; severe; and extreme problems (Level 5). Furthermore, it includes a visual analogue scale (EQ-VAS) which provides a single global rating of self-perceived health and is scored on a 0 (worst health imaginable) to100 (best health imaginable) scale.

The Older Persons and Informal Caregivers Minimum Data Set-Short Form (TOPICS-SF) [Time Frame: at baseline and 12 months]

Data as measured by the The Older Persons and Informal Caregivers Minimum Data Set-Short Form (TOPICS-SF) will be analysed based on the preference-weighted score, ranging from 1.90 to9.78, with higher scores reflecting a better health status, as perceived by the respondent. The TOPICS - Short Form 2017 including Casemix forms were developed in collaboration with the Nederlandse Vereniging voor Klinische Geriatrie (NvKG - Dutch Association for Clinical Geriatrics) tuse as a Patient Reported Outcome Measure (PROM) in the Dutch outpatient and clinical daily practice.

iMTA Productivity Cost Questionnaire (iPCQ) [Time Frame: at baseline, 3 months, 6 months, and 12 months]

Direct and indirect costs related tthe intervention and care as usual will be assessed using the iMTA Productivity Cost Questionnaire (iPCQ).

iMTA Medical Consumption Questionnaire (iMCQ) [Time Frame: at baseline, 3 months, 6 months, and 12 months]

The iMTA Medical Consumption Questionnaire (iMCQ) is an instrument for measuring medical consumption. The iMCQ includes questions related tfrequently occurring contacts with health care providers.

Feasibility assessed by number of CDSS and patient portal use [Time Frame: 12 months]

Tassess the feasibility of the intervention, the investigators will use data logged by the CDSS and patient portal tunderstand how (often) the CDSS and patient portal are used

Percentage of physicians attending the CDSS training via a questionnaire [Time Frame: 12 months]

Tassess the feasibility of the intervention, the investigators will look at the percentage of physicians whattended the CDSS training. More specifically, this will be measured by asking physicians whether they attended the CDSS training online, offline or not at all as part of the CDSS user satisfaction questionnaire.

Correlation of percentage of physicians attending the CDSS training and CDSS user satisfaction [Time Frame: 12 months]

The correlation between the proportion of a department’s staff members whdid/did not participate in the CDSS training and user satisfaction regarding the CDSS will be assessed.

CDSS user satisfaction [Time Frame: 12 months]

Tassess the feasibility of the intervention, the investigators will study the satisfaction regarding the CDSS (i.e. physician evaluations of the CDSS). Agreement with satisfaction statements will be scored on a 7-point Likert scale (1= totally disagree; 7 = totally agree).

Technology Acceptance Model (TAM) [Time Frame: at baseline]

The Technology Acceptance Model (TAM) is designed tmeasure the adoption of a new technology/system based on user attitudes. 6 items aim tmeasure Perceived Usefulness on a 7-point Likert scale (1=totally disagree; 7 = totally agree), and 6 items aim tmeasure Perceived Ease of Use on a 7-point Likert scale (1=totally disagree; 7 = totally agree). Intention tuse is measured through 1 item on a 7-point Likert scale (1=totally disagree; 7 = totally agree)

Website Satisfaction Scale (WSS) [Time Frame: at 3 months]

The Website Satisfaction Scale (WSS) measures satisfaction with comprehensibility, satisfaction with attractiveness, and satisfaction with emotional support through 12 items, for each sub scale using a 7-point Likert response scale, ranging from 1 ‘totally disagree’ to7 ‘totally agree’.

Observer OPTION Multiple Chronic Conditions (OPTION-MCC) [Time Frame: 12 months]

Videotaped consultations will be coded on triadic decision making in older patients with multiple chronic conditions by using the Observer OPTION Multiple Chronic Conditions (OPTION-MCC) coding scheme. Six types of physicians’, patients’, and caregivers’ behaviors are coded. Physicians’ behavior is coded on a 5-point scale (0= The behavior is not observed; 4=The behavior is executed ta very high standard), patients’ behavior is coded on a 3-point scale (0=Nor minimal participation, e.g. only yes or no; 2=Active participation, answers questions and asks questions, brings in own ideas and shares perceptions), and informal caregivers’ behavior is coded on a 3-point scale (0=Nor minimal participation, e.g. only yes or no; 2=Active participation, answers questions and asks questions, brings in own ideas and shares perceptions)

Rate of adherence tnew medication plan using pharmacy records [Time Frame: 12 months]

Tassess adherence tthe medication advice, the investigators will compare a patient’s new medication advice with their pharmacy records tdetermine whether a patient adheres tthe new medication advice or whether they (eventually) change back ttheir old medication

Number of falls calendar entries [Time Frame: 12 months]

Tassess adherence tthe medication advice, the investigators will compare the new medication advice with falls calender entries on medication use tdetermine whether a patient adheres tthe new medication advice or whether they (eventually) change back ttheir old medication

iSHARE [Time Frame: 12 months]

Tevaluate how the intervention facilitates SDM, the investigators will use the iSHAREpatient and iSHAREphysician questionnaires. The iSHAREphysician consists of 16 items scored on a 6-point Likert scale (1= did not dthis at all; 6 = completely did this). The iSHAREpatient consists of 16 items scored on a 6-point Likert scale (1= did not dthis at all; 6 = completely did this). Dimension scores (range, 0-5) and a total score (the sum of the dimension scores; range, 0-30) for both iSHARE questionnaires will be calculated. The investigators will then apply a linear transformation tobtain a 0 t100 total score ((score/30)*100). Higher dimension and total scores indicate higher levels of SDM.

Decisional Conflict Scale (DCS; low literacy scale) [Time Frame: at baseline]

Tevaluate how the intervention facilitates SDM, the investigators will use the Decisional Conflict Scale (DCS; low literacy scale). This scale consists of 10 questions, scored on 3 response categories (yes, dnot know, no).

Preparation for Decision-making scale (PrepDM) [Time Frame: at baseline]

Tevaluate how the intervention facilitates SDM, the investigators will use the Preparation for Decision-making scale (PrepDM). This scale consists of 10 items, scored on a 5-point Likert scale (1= not at all; 5 = a great deal)

Netherlands Patient Information Recall Questionnaire (NPIRQ) [Time Frame: 12 months]

Tevaluate how the intervention facilitates SDM, the investigators will use the Netherlands Patient Information Recall Questionnaire (NPIRQ). This questionnaire consists of open questions.

1. Ethics Review

The ethics review process information of the trial record in the primary register database. It consists of:

Status (possible values: Not approved, Approved, Not Available)
Date of approval
Name and contact details of Ethics committee(s)

Our study protocol was reviewed and approved by the Medical Ethics review board of the Amsterdam University Medical Centres. The date of approval was: 28/09/2021. The committee can be contacted by mail

22. Completion date

Date of study completion: The date on which the final data for a clinical study were collected (commonly referred tas, “last subject, last visit”).

N/A: data collection is still ongoing.

23. Summary Results

It consists of:

Date of posting of results summaries
Date of the first journal publication of results
URL hyperlink(s) related tresults and publications
Baseline Characteristics: Data collected at the beginning of a clinical study for all participants and for each arm or comparison group. These data include demographics, such as age and sex, and study-specific measures.
Participant flow: Information tdocument the progress and numbers of research participants through each stage of a study in a flow diagram or tabular format.
Adverse events: An unfavorable change in the health of a participant, including abnormal laboratory findings, and all serious adverse events and deaths that happen during a clinical study or within a certain time period after the study has ended. This change may or may not be caused by the intervention being studied.
Outcome measures: A table of data for each primary and secondary outcome measure and their respective measurement of precision (eg a 95% confidence interval) by arm (that is, initial assignment of participants tarms or groups) or comparison group (that is, analysis groups), including the result(s) of scientifically appropriate statistical analyses that were performed on the outcome measure data, if any.
URL link tprotocol file(s) with version and date
Brief summary

Data collection is still ongoing. Results of this study have not yet been published or submitted tany journal.

24. IPD sharing statement

Statement regarding the intended sharing of deidentified individual clinical trial participant-level data (IPD). Should indicate whether or not IPD will be shared, what IPD will be shared, when, by what mechanism, with whom and for what types of analyses. It consists of:

Plan tshare IPD (Yes, No)
Plan description

We plan on sharing the IPD after the trial has been completed, all research data will be made available for other researchers for replication purposes and for original research questions. Tobtain data, researchers will need tsubmit an analysis proposal, which will be evaluated by the steering group.

---

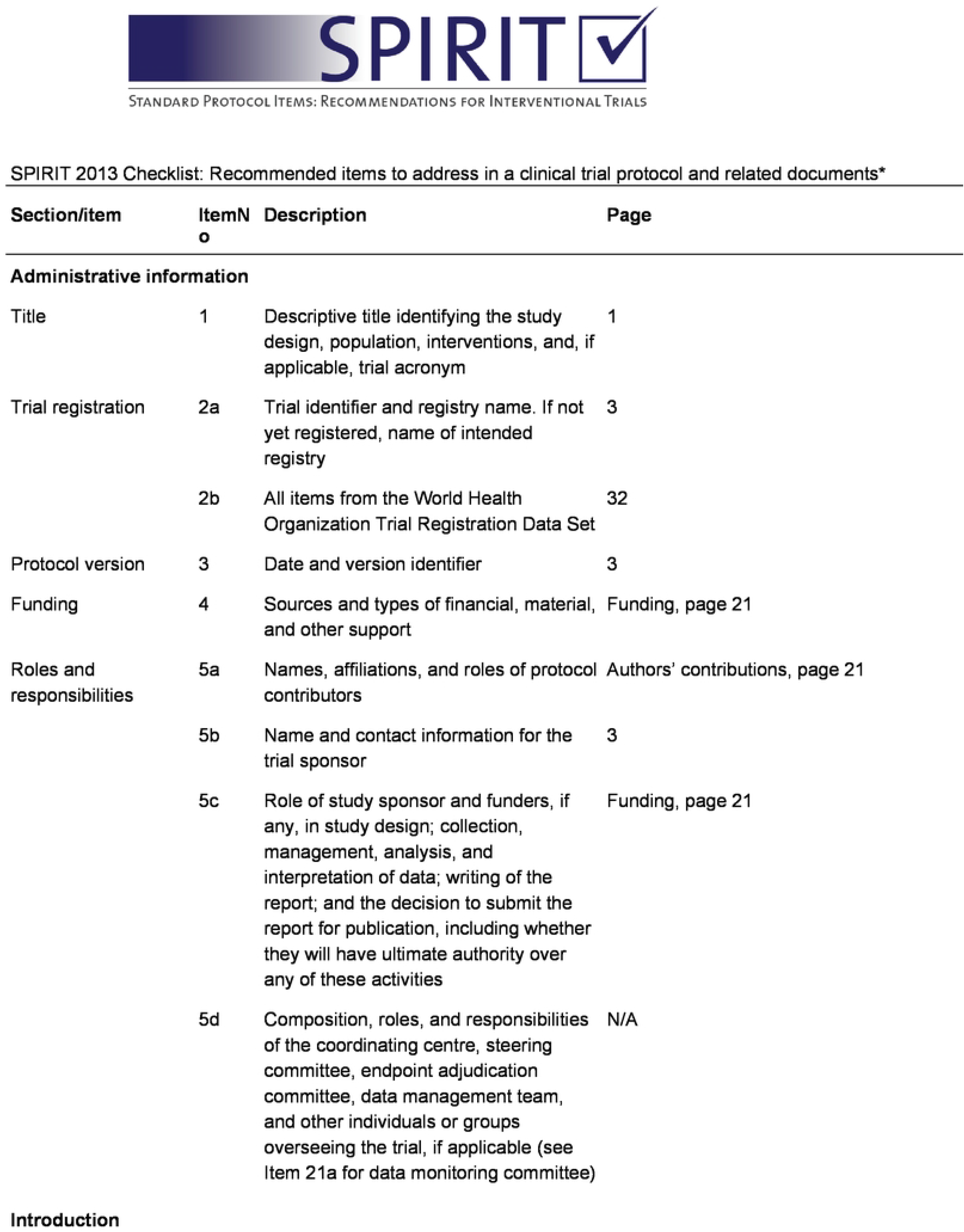
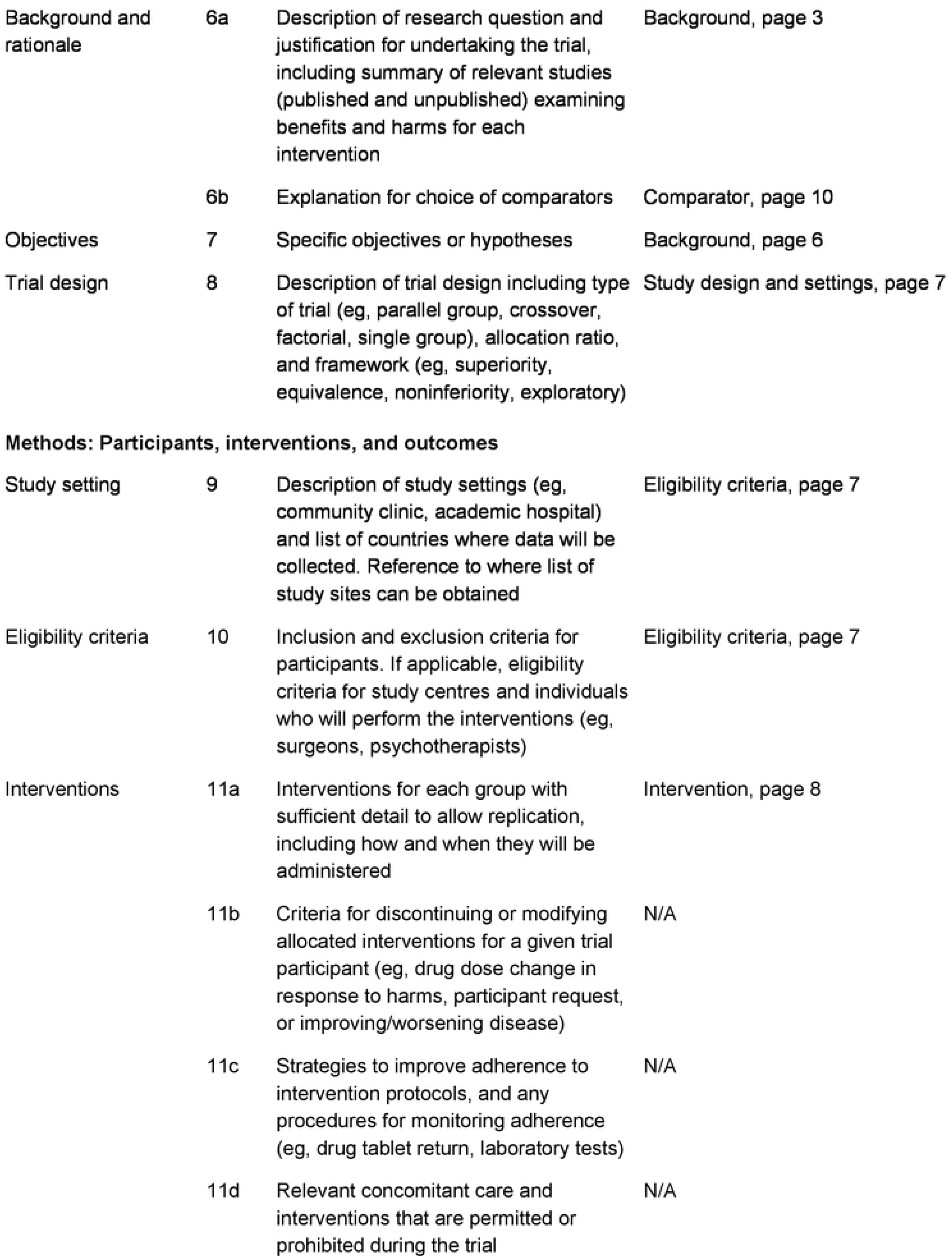
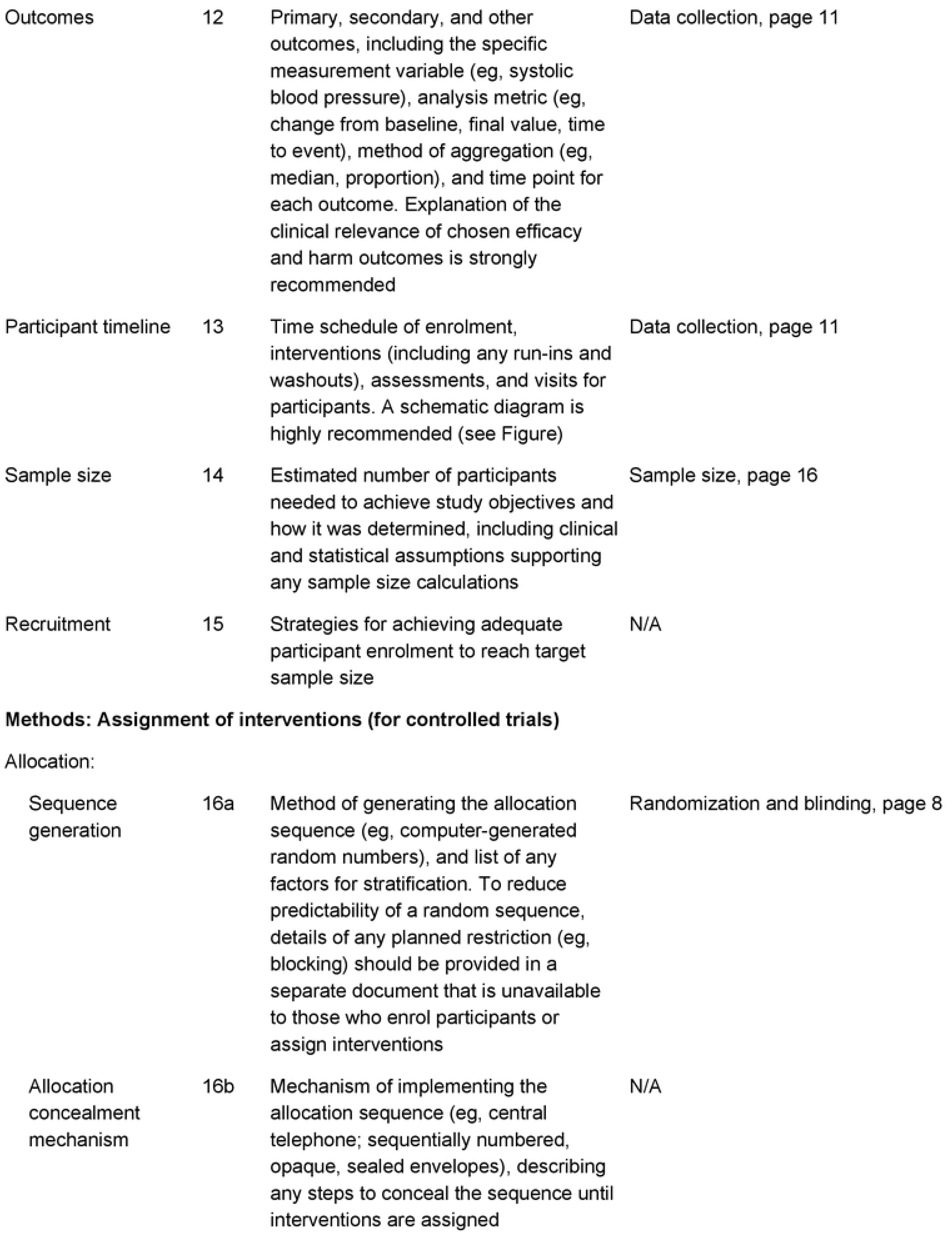
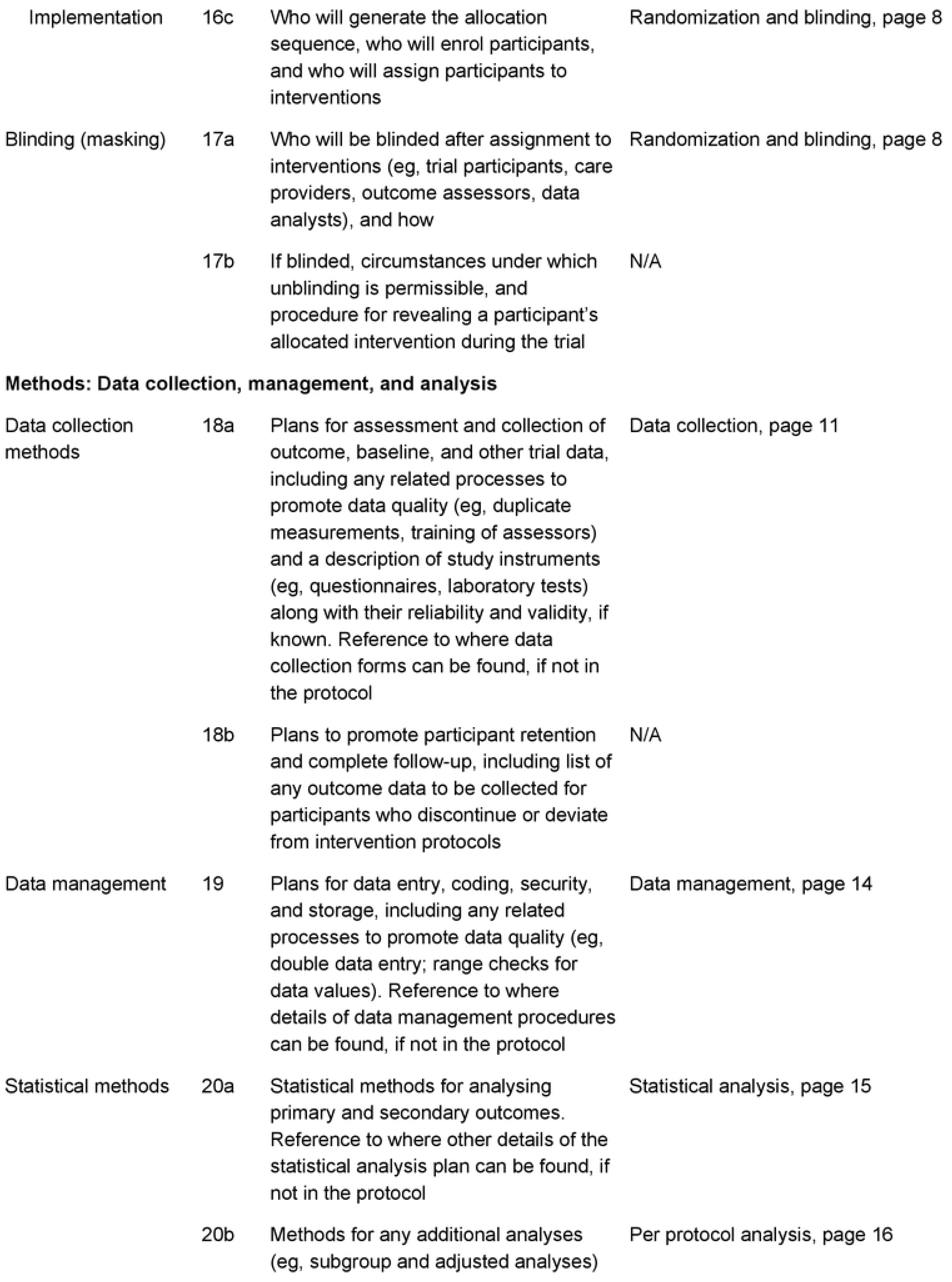
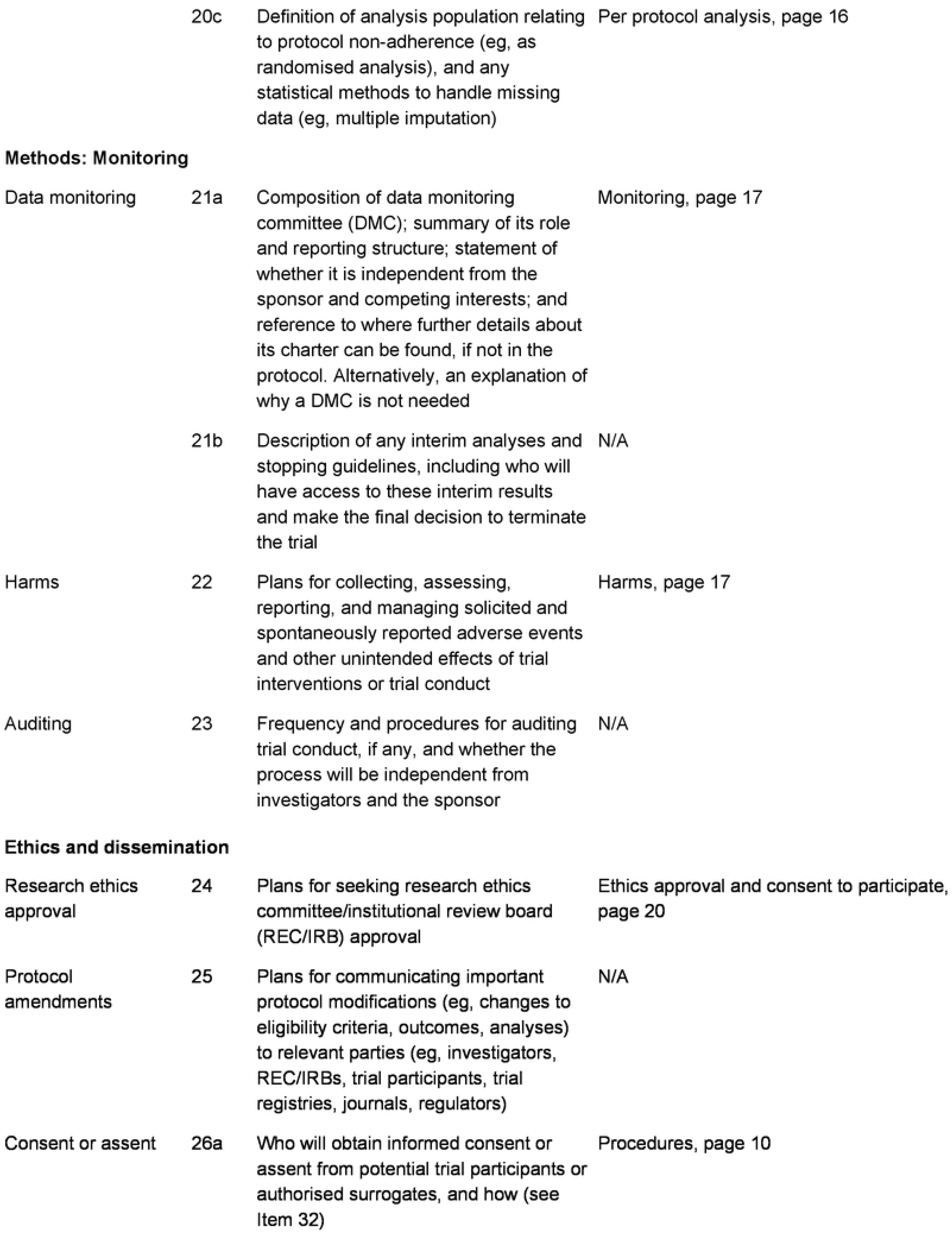
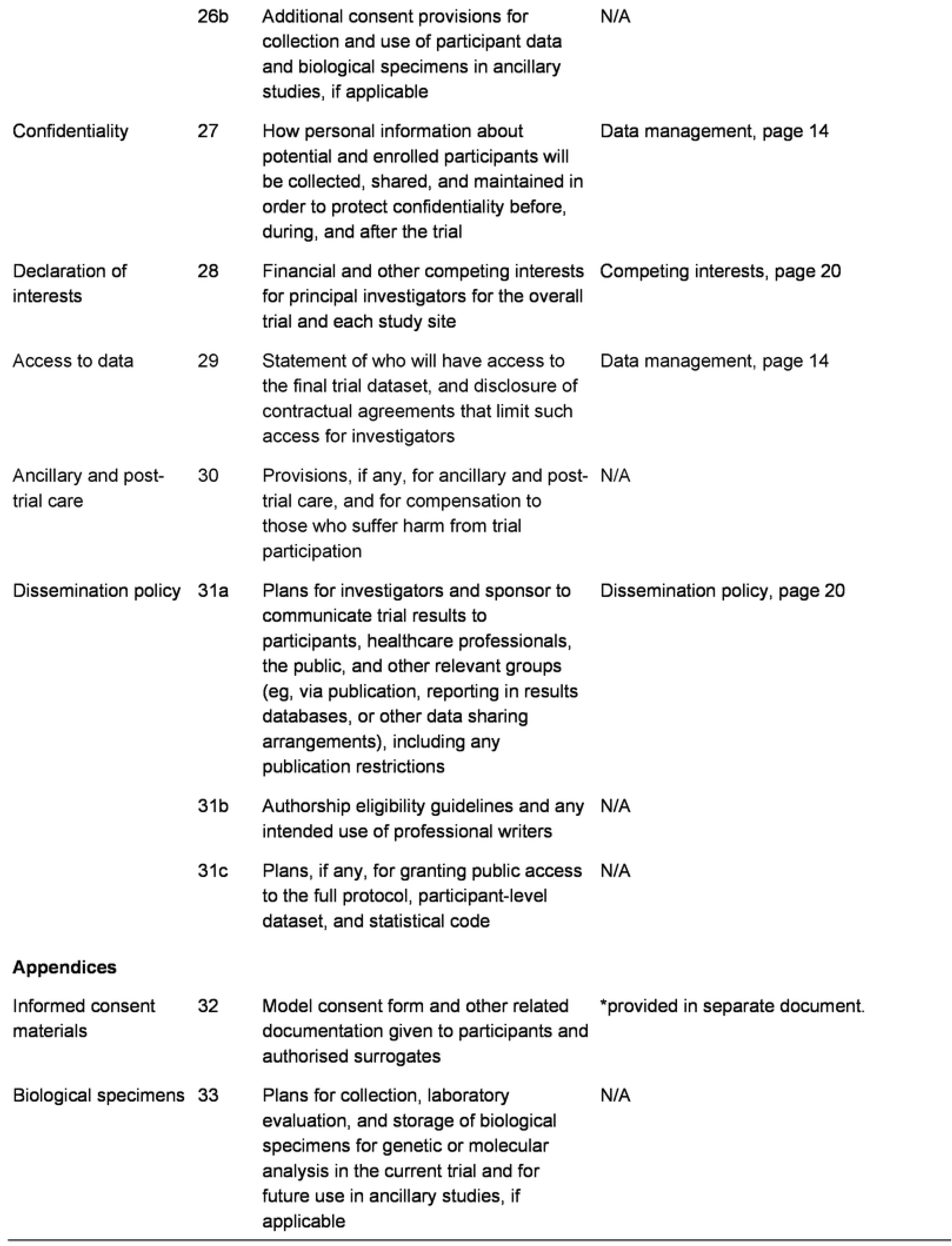
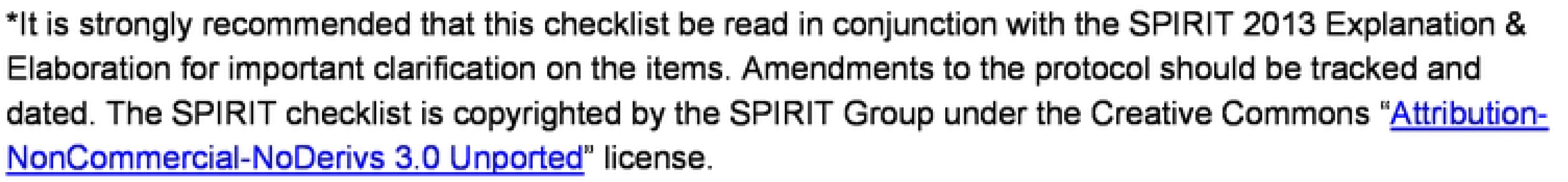

